# Psychological distress after COVID-19 recovery and subsequent prolonged post-acute sequelae of COVID-19: A longitudinal study with 1-year follow-up in Japan

**DOI:** 10.1101/2024.03.21.24304702

**Authors:** Megumi Hazumi, Mayumi Kataoka, Zui Narita, Kentaro Usuda, Emi Okazaki, Daisuke Nishi

## Abstract

**Background:** This study investigated the longitudinal association between psychological distress in the post-acute phase and the subsequent prolonged post-acute sequelae of COVID-19 (PASC) among individuals with PASC.

**Methods:** An online longitudinal survey with 1-year interval was conducted from July to September 2021 (T1) and July to September 2022 (T2). Individuals who were 20-years-old or older, had a positive Polymerase Chain Reaction test, were one month post-infection, and did not select “Nothing” to a question regarding PASC presence were included. The primary outcome was the presence of PASC at T2. The presence of general, respiratory, muscular, neurological, gastrointestinal, dermatological, and cardiac symptoms at T2 was also used as an outcome among patients with relevant symptoms at time 1 (T1). Exposure was measured using the Kessler distress scale (K6) at T1, and those whose K6 was 13 or higher were identified as having psychological distress. Marginal structure models with robust standard errors were used to examine the association between psychological distress at T1 and any PASC symptoms at T2, and the associations between psychological distress and each symptom at T2 among participants with relevant symptoms at T1.

**Results:** A total of 1674 patients were analyzed; 17%had psychological distress. ; In total, 818 (48.9%), 523 (31.2%), and 672 (40.1%) patients reported general, respiratory, and neurological symptoms at T1, respectively. Individuals with psychological distress had higher odds of any symptoms at T2 (Odds Ratio [OR] =1.81, 95% Confidence Interval [CI]= 1.08 – 3.03) and general and respiratory symptoms at T2 among participants with relevant symptoms at T1 (OR = 1.95, 95% CI = 1.02 – 3.76; OR = 2.44, 95% CI = 1.03 – 5.80).

**Conclusion:** Psychological distress in the post-acute phase may lead to prolonged PASC symptoms, mainly general and respiratory symptoms, at the 1-year follow-up in individuals with PASC.

**Key Messages:** Psychological distress before infection and during the acute phase predicts prolonged Post-Acute Sequelae of COVID-19 (PASC); however, in individuals with PASC, it is unclear whether psychological distress during the post-acute phase predicts prolonged PASC. This longitudinal survey indicated that psychological distress in the post-acute phase led to prolonged any levels of PASC, especially general and respiratory symptoms, at the 1-year follow-up. Therefore, mental health care for individuals with PASC may help to improve or mitigate prolonged PASC.

## Introduction

Post-acute sequelae of COVID-19 (PASC) are symptoms that persist or appear after the acute phase, that is, after four weeks of COVID-19 infection[1]. PASC symptoms vary, but may include dyspnea, cough, fatigue, fever, anosmia, ageusia, arthralgia, myalgia, hair loss, chest pain, palpitations, throat pain, and others[2,3]. PASC can last more than several months and is as severe as other infectious diseases. According to a systematic review, 54%, 55%, and 54% of COVID-19 survivors had at least one PASC at one month, two to five months, and six or more months, respectively[1], and 50.1% continued to suffer from PASC for more than one year[4]. Abnormal pulmonary function (45.6%), fatigue (28.7%), and neurological symptoms (18.7%) were mainly observed[4]. The prevalence of symptoms was significantly higher in individuals infected with COVID-19 than in those without after one year had passed[5], and such symptoms were not significantly reduced between one year and two years after infection[6]. The prevalence of sequelae in patients with COVID-19 is comparable to that in other respiratory infectious diseases[7]. Considering that persistent symptoms not only affect these individuals’ quality of life[8,9] but are also costly from an economic perspective[10–12], it is important to identify factors that cause PASC to persist so that it can be alleviated and prevented.

Having mental health problems before COVID-19 infection and in the acute phase is suggested to be one factor that can cause prolonged PASC. The association between preexisting mental health problems and PASC has been reported in several cross-sectional surveys[13–17]. According to several longitudinal studies, preexisting mental health problems predict PASC at 3, 12, and 15 months after infection[15,18–20]. An extensive cohort study reported that pre-existing depression predicted prolonged PASC after 6, 12, and 18 months passed[21]. Mental health problems in the acute phase have also been suggested to influence PASC. A prospective cohort study indicated that depressive symptoms in the acute phase predicted the presence of PASC at one and three months after infection [22]. Another prospective cohort study revealed that depression in the acute phase predicted prolonged PASC at 12 months after infection[23]. Considering these factors, mental health problems before the appearance of PASC may determine whether PASC will persist for more than 12 months.

However, it remains unclear whether mental health problems after the appearance of PASC predict prolonged PASC. Several studies suggest that mental health problems may develop or continue after the acute phase among those with PASC. Individuals who recover from COVID-19 are at risk of developing new mental health problems[4,24–27]. COVID-19-related experiences, such as perceived discrimination and negative thoughts about the infection, were reported to be associated with mental health problems, independent of pre-existing mental health problems[28,29]. Several cross-sectional studies also suggest that preceding PASC affects mental health after recovery[30–34], and longitudinal studies have suggested that presence of PASC predicts mental health problems[35,36]. On the other hand, mental health problems after acute illness are suspected to predict subsequent prolonged PASC, considering that mental health problems generally exacerbate the severity or progression of various physical diseases[37]. Psychological distress after the appearance of to contribute to prolonged PASC as it is possible for mental health to develop after the acute phase of infection, and the possibility of mental health problems exacerbating physical health problems. Various PASC symptoms, both general symptoms, such as fatigue and fever, and respiratory symptoms, including dyspnea and shortness of breath, may be influenced by mental health problems. The association between respiratory symptoms and mental health problems is well known[38,39] because negative feelings generally promote the perception of dyspnea, airway obstruction, and airway reactivity through the CNS and autonomic pathways[40,41]. General symptoms, such as fatigue and fever, are generally affected by mental health problems[42–44] through the elevation of interleukin-1 (IL-1) and IL-6, which induce fatigue and fever[45,46].

Therefore, this study aimed to investigate whether psychological distress after the acute phase predicts subsequent prolonged PASC at a one year follow-up afting forunder adjusted pre-existing mental health problems in individuals with PASC. This study is believed to help elucidate the significance of providing mental health care to individuals with PASC with the aim of alleviating their symptoms.

## Materials and Methods

### Participants and Setting

The two-point longitudinal survey for COVID-19 was performed at 1-year intervals through the Rakuten Insight Corporation, which is the company that delivering the most mass panels in Japan for online surveys[47]. The panelists received information about study recruitment, and those interested in the study accessed an online survey form. If the individuals agreed to participate in the study by clicking the consent declaration button on the website after reading the informed consent document, they were considered to have responded to the questionnaire. Those who completed the questionnaire received rewards points that could can be used for online shopping. The data at Time 1 (T1) and Time 2 (T2) were collected from July to September 2021 and July to September 2022, respectively.

Individuals who met the following criteria were included: 1) Over 20 years old, 2) answered “yes” to the screening question “Have you been infected with COVID-19?”, 3) selected an option other than “Nothing” to the question “What kind of PASC symptoms do you have now?”, and 4) more than a month after infection, based on the definition of PASC[48]. In addition, data that met the following criteria were excluded from the analysis: duplication, incorrect answers to the dummy question, statement that they had not been infected, inconsistent answers, and outlier answers according to their demographic information responses.

### Measurements

#### Outcome

The answer options to the question “What kind of physical PASC symptoms do you have now?” at T2 were used as the outcomes. Options were generated based on the symptoms list by the World Health Organization[49] and the detailed answers of “Others” symptoms at T1: “Nothing,” “Fever,” “Cough,” “Fatigue,” “muscle or body pain,” “Sore throat,” “Headache,” “Diarrhea,” “Red or irritated eyes,” “Altered taste,” “Altered smell,” “A rash on skin, or discoloration of fingers or toes,” “Difficulty breathing or shortness of breath,” “Chest pain or pressure,” “Loss of speech or mobility,” “Menstrual problems,” “Blurred vision,” “Dizziness,” “Constipation or acid reflux” “Convulsion,” “Neuralagia,” “Tachysystole and palpitation,” “Tinnitus and ear problems,” “New Allergy” and “Others.” Unless the respondent selected the “nothing” option, they were able to select multiple options. Those who selected “Others” were additionally asked to fill in the details in the free text field, and authors categorized the described complaint by compiling similar symptoms. Furthermore, these symptoms were classified into the following symptom groups based on a previous study[50]: “General,” “Respiratory,” “Musculoskeletal,” “Neurologic,” “Gastrointestinal,” “Eye,” “Dermatologic,” “Cardiac,” “Urinary,” and “Others.” Details regarding the symptoms and classifications are provided in Appendix 1.

To confirm whether any physical PASC was maintained, the option “Nothing” at T2 was used as a primary outcome. Those who selected “Nothing” were identified as having no PASC, and those who didn’t were identified as having some PASC. The presence of each symptom group at T2 was also used as an outcome measure.

These variables and classification procedures were also employed for the data at T1 in subgroup analyses.

#### Exposure

The Kessler Psychological Distress Scale (K6) at T1 was used to measure the severity of psychological distress over the past 30 days at T1[51–53]. The K6 comprises six items with 5-point Likert scales (0–4 points). A total score of 13 or higher indicates psychological distress[51].

#### Covariates

The following variables reported at T1 were used as covariates: sex (male, female, others)[54,55], age group (20–29, 30–39, 40–49, 50–59, ≥ 60)[54,55], income level (< Japanese yen [JPY] 3,000,000, < JPY 10,000,000, ≥ JPY 10,000,000, unknown or refuse to respond)[56,57], the presence of psychiatric history (yes, no)[24,54,55], hospitalized (yes, no)[54,58], and the presence of acute symptoms (yes, no)[55,58]. We also adjusted for BMI[48,54,59] and pre-existing comorbidities, including respiratory diseases (COPD, asthma, bronchitis, and pneumonia)[54,55,60], hypertension[55,61,62], diabetes[48,59,63], and cardiac diseases (angina and myocardial infarction)[48,64]. These pre-existing comorbidities were identified as those who selected options other than “Nothing before” or “Never,” “Yes, in the past,” “Yes, currently (in hospitals),” and “Yes, currently (not in hospitals)” to the question about above comorbidities.

### Patent and Public Involvement

From inception, our research team included COVID-19 survivors to leverage their first-hand experiences in developing the hypothesis and ensuring patient-centered perspectives were integral to our study design.

### Analyses

After absolute risks and risk differences were calculated, marginal structure models with robust standard errors were performed to examine the association between the presence of psychological distress at T1 and the scprecense of PASC symptoms at T2[65]. Similar analyses were performed to examine the association between the presence of psychological distress at T1 and the presence of each symptom at T2, including “General,” “Respiratory,” and “Neurologic,” among those with relevant symptoms at T1, given that sample size at T1 was sufficient. covariates were controlled to use inverse probability weighting. Stabilized weights were calculated to determine the presence of psychological distress. We further accounted for the loss to follow-up using stabilized weights. The stabilized weights were multiplied to obtain the final stabilized weights.

### Ethics

This study was performed in accordance with the Declaration of Helsinki and was approved by the Research Ethics Committee of the National Center of Neurology and Psychiatry (A2021-034).

## Results

### Characteristics

Figure 1 illustrates the data collection process. A total of 1674 eligible participants completed the questionnaire at T1, and their data were analyzed. At T2, 673 participants completed the questionnaire.

Table 1 presents the demographic characteristics of the participants. Of all participants, 57.6% were male, and the age group of 40–49 years was the largest (29.2%). The proportion of those whose have psychological distress was 17%.

**Table 1.** Characteristics.

|  | n=1674 |  |
| --- | --- | --- |
|  | n/ mean | %/ SD |
| Sex |  |  |
| Male | 964 | 57.6% |
| Female | 699 | 41.8% |
| Other | 11 | 0.7% |
| Age |  |  |
| 20 - 29 | 271 | 16.2% |
| 30 - 39 | 407 | 24.3% |
| 40 - 49 | 488 | 29.2% |
| 50 - 59 | 371 | 22.2% |
| ≥ 60 | 137 | 8.2% |
| Income |  |  |
| < JPY3,000,000 | 247 | 14.8% |
| < JPY10,000,000 | 1031 | 61.6% |
| ≥ JPY10,000,000 | 249 | 14.9% |
| Unknown or refuse to respond | 147 |  |
| BMI | 24.01 | 4.42 |
| Physical comorbidity |  |  |
| Hypertension | 469 | 28.0% |
| Diabetes | 238 | 14.2% |
| Respiratory diseases | 651 | 38.9% |
| Cardiac diseases | 155 | 9.3% |
| Psychiatric history | 478 | 28.6% |
| Hospitalized | 639 | 38.2% |
| Duration after infection (month) | 5.12 | 4.25 |
| Any acute symptoms | 1665 | 99.5% |
| Psychological distress (K6 ≥ 13) | 284 | 17.0% |
| PASC symptom groups |  |  |
| General | 818 | 48.9% |
| Respiratory | 523 | 31.2% |
| Musculoskeletal | 190 | 11.4% |
| Neurologic | 672 | 40.1% |
| Gastrointestinal | 72 | 4.3% |
| Eye | 24 | 1.4% |
| Dermatologic | 137 | 8.2% |
| Cardiac | 143 | 8.5% |
| Urinary | 2 | 0.1% |
| Other | 23 | 1.4% |

PASC, Post-acute sequelae of COVID-19; BMI, Body Mass Index; JPY, Japanese Yen; K6, Kessler distress scale

### Relationships between psychological distress and subsequent prolonged PASC

Table 2 shows the absolute risks and risk differences of the presence of prolonged PASC between those with psycological distress and those without.

**Table 2.** Absolute risks and risk differences of PASC at 1-year follow-up.

|  | Psychological distress |  | Risk difference |
| --- | --- | --- | --- |
|  | Yes | No |  |
| Any symptoms | 66.1% | 50.7% | 15.3% |
| General | 55.9% | 34.3% | 21.5% |
| Respiratory | 51.3% | 32.4% | 18.9% |
| Neurologic | 47.3% | 43.0% | 4.3% |

As table 3 shows, psychological distress at T1 was significantly associated with the presence of any symptoms, general symptoms, and respiratory symptoms at T2 among those with the relevant symptoms at T1 (OR=1.80 [1.08 – 3.01]; OR=1.92[1.002 – 3.67]; OR=2.44[1.03 – 5.80]).

**Table 3.** Psychological distress as a predictor of prolonged PASC at 1-year follow-up.

|  | 95% CI |  |  |
| --- | --- | --- | --- |
|  | OR | Lwr | Upr |
| Any symptoms | 1.81 | 1.08 | 3.03 |
| General | 1.95 | 1.02 | 3.76 |
| Respiratory | 2.44 | 1.03 | 5.80 |
| Neurologic | 1.14 | 0.58 | 2.26 |

OR = Odds Ratio; 95% CI = 95% confidence interval; Lwr, Lower; Upr, Upper

## Discussion

This study investigated the association between psychological distress after recovery from COVID-19 and PASC at one-year follow-up in individuals with PASC after COVID-19 infection. Psychological distress after recovery was significantly associated with subsequent proalonged PASC, both general and respiratory symptoms, at one year among those with relevant symptoms, independent of pre-existing psychiatric problems.

These results suggested that psychological distress at the post-acute phase wasassociated with prolonged PASC of any type. Inaddition to the mental health problems observed before the post-acute phase[15,18–20,22,23,66], psychological distress after PASC becomes apparent may lead to prolonged PASC. COVID-19 survivors often face discrimination, negative thoughts about COVID-19, and experience unemployment after recovery, which can exacerbate mental health problems [28,29][67][68]. These factors may, in turn, lead to prolonged PASC due to resultant mental health problems.

Psychological distress after the acute phase is associated with prolonged general symptoms, such as fatigue and fever, in individuals with PASC. This result is consistent with general symptoms not induced by COVID-19 infection[42–44]. The depressive conditions are known to elevate IL-1β and IL-6, which cause fever and fatigue[45,46]. Mental health problems related to PASC and COVID-19 are believed to activate inflammatory cytokines such as IL-1β and IL-6 that can prolong general symptoms.

Psychological distress after the acute phase is associated with prolonged respiratory symptoms, such as dyspnea and shortness of breath. As respiratory symptoms are unrelated to COVID-19 infection[38,39], respiratory symptoms, such as PASC, may be affected by mental health problems.

Respiratory symptoms, such as dyspnea, obstruction, and airway sensations, are sensitive to negative emotions[40,41]. Therefore, mental health problems may promote the perception of respiratory symptoms and delay improvements in PASC.

Psychological distress was not associated with prolong neurological symptoms, supporting previous findings that the oresence of psychological distress did not differ significantly between those with edaltering smell or taste after COVID-19 infection and those without [13].

These findings underscore the need to routinely assess mental health as a critical aspect of PASC care. Early identification and support for psychological distress may reduce the risk of prolonged PASC. These implications are not limited to the clinical realm for medical professionals but also extended to larger scales, such as regional and societal levels, indicating significant public health implications.

This study had some limitations. PASC symptoms were self-reported by the participants. Thus, they were not as subtle as diagnoses made by physicians or determined using validated measurements. Instead, our findings reflect individuals with PASC who are not connected to medical care. We failed to follow-up with over half of the participants, although missing values were complemented by a statistical procedure to minimize missing bias. It cannot be denied that the mental health problems measured in this study emerged prior to the PASC, although mental health problems before the infection were adjusted. The results for several symptoms derived from the small sample size should be interpreted carefully.

There are also some strengths to this study. Our data included individuals who were and were not seeing doctors. As some individuals with PASC find it challenging to access healthcare[69,70], collecting participants from both groups is believed to reflect comprehensive PASC characteristics.

In conclusion, this study revealed the relationship between mental health problems in the post-acute phase and subsequent prolonged PASC at the 1-year follow-up in individuals with PASC. General and respiratory symptoms were associated with mental health problems. Mental care may reduce the incidence of PASC, especially general and respiratory symptoms. Future intervention studies targeting mental health problems related to PASC should be conducted.

## Data Availability

Data produced in the present study are not available.

## Contributorship statement

Conceptualization: all. Data collection: MH, MK, KU, EO, and DN. Formal analysis: MH. Interpretation of data: MH, MK, ZN and DN. Original Draft preparation; MH. Review and editing: all. Super vision: ZN and DN.

## Appendix 1.

The symptom classification

Appendix 1. The symptom classification
| Symptom group | Classified symptoms |
| --- | --- |
| **General** | Fever, Fatigue, Loss of speech or mobility, Rigor*, Sweat*, Edma and swell* |
| **Respiratory** | Cough, Sore throat, Difficulty breathing or shortness of breath, Other throat problems* |
| **Musculoskeletal** | Muscle or body pain |
| **Neurologic** | Headache, Altered taste, Altered smell, Dizziness, Convulsion, Neuralgia, Tinnitus and ear problems |
| **Gastrointestinal** | Diarrhea, Constipation or acid reflux, Anorexia* |
| **Eye** | Red or irritated eyes, Blurred vision |
| **Dermatologic** | A rash on skin, or discoloration of fingers or toes, Hair loss* |
| **Cardiac** | Chest pain or pressure, Tachysystole and palpitation |
| **Urinary** | Urinary* |
| **Others** | Menstrual problems, New allergy, other complaints |

\*categories generated based on the detailed description of the alternative “Others”

The categories “Ear,” “Metabolic,” and “Reproductive” were not developed because the responses equivalent to these categories were not collected.

## Figure Captions

Figure 1. Flow chart

